# Synchronization in Epidemic Growth and the Impossibility of Selective Containment

**DOI:** 10.1101/2020.11.06.20226894

**Authors:** Jan Carl Budich, Emil J. Bergholtz

**Affiliations:** Institute of Theoretical Physics, Technische Universität Dresden and Würzburg-Dresden Cluster of Excellence ct.qmat, 01062 Dresden, Germany; Department of Physics, Stockholm University, AlbaNova University Center, 106 91 Stockholm, Sweden

## Abstract

Containment, aiming to prevent the epidemic stage of community-spreading altogether, and mitigation, aiming to merely ‘flatten the curve’ of a wide-ranged outbreak, constitute two qualitatively different approaches to combating an epidemic through non-pharmaceutical interventions. Here, we study a simple model of epidemic dynamics separating the population into two groups, namely a low-risk group and a high-risk group, for which different strategies are pursued. Due to synchronization effects, we find that maintaining a slower epidemic growth behavior for the high-risk group is unstable against any finite coupling between the two groups. More precisely, the density of infected individuals in the two groups qualitatively evolves very similarly, apart from a small time delay and an overall scaling factor quantifying the coupling between the groups. Hence, selective containment of the epidemic in a targeted (high-risk) group is practically impossible whenever the surrounding society implements a mitigated community-spreading. We relate our general findings to the ongoing COVID-19 pandemic.

---

The ongoing COVID-19 pandemic caused by the new Coronavirus SARS-CoV-2 is among the biggest global challenges of our time [1], and its quantitative analysis has thus been an intense focus of recent research [2–7]. A repeatedly debated [8, 9] mitigation strategy is based on selectively protecting vulnerable individuals that are at high risk to die or at least develop a severe condition when contracting the virus. Such a strategy, in the following referred to as 2GROUPS, in practice amounts to defining (at least) two groups of individuals, a low-risk group (L) and a high-risk group (H), and then focusing most of the available resources to try and protect the group H from infection, while the larger low-risk group basically does “business as usual”, possibly combined with moderate general mitigation measures aimed at ‘flattening the curve’ of infections for group L.

Here, we analyze the expected qualitative outcome of such a 2GROUPS scenario [10]. Generally speaking, there are two crucial ingredients to a 2GROUPS strategy: First, efficient criteria to identify the high-risk and the low-risk individuals a priori. Regarding this aspect, earlier studies indicated that a large fraction of anticipated severe cases may be concentrated in a relatively small group H, even if only the single criterion of age is used [2, 11]. Second, the isolation of the high-risk group aimed at strongly containing the prevalence of the disease in group H. In this regard, we find that the selective isolation of the risk group is largely stymied by synchronization effects in the equations governing the pandemic dynamics (see Fig. 1 for an illustration). In particular, we analytically demonstrate within a minimal mathematical model that maintaining a qualitatively slower growth of infections in the high-risk group is unstable against any finite coupling between the two groups, and thus impractical. Instead, the overall exposure of the group H is found to be proportional to that of group L, which may allow for a certain degree of mitigation within H, but largely rules out selective containment within the high-risk group.

**FIG. 1:**
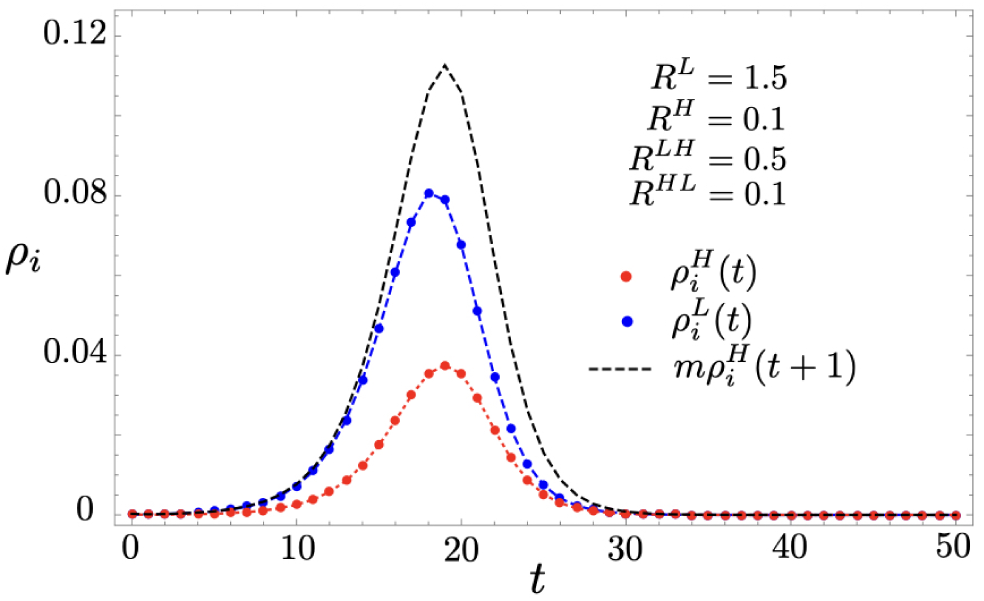
Syncronization of epidemic growth governed by Eq. 1. Even if the high-risk group (H) isolates very efficiently, here quantified by a small basic reproduction number 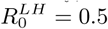, a significant fraction 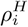 (red) becomes infected if the low-risk group (L) exhibits a mitigated ‘flatten the curve’ scenario, here at 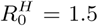, as described by 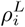 (blue). We show that this holds for any finite coupling between the groups due to a synchronization effect taking place in exponential growth phase of the epidemic. Specifically, during the synchronization the high and low-risk group infection rates are scaled by a simple constant factor, 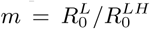 and shifted by one time-step (cf. the black dashed curve). After the synchronization the infection rate in the high-risk group remains significant and bounded from below as 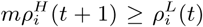. Initial conditions set to 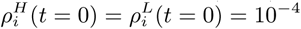.

## Minimal mathematical model of 2GROUPS

We analyze a simple two-component model of SIR type [12], in which the qualitative synchronization between the epidemic curves of the two groups (cf. Fig. 1) may be readily understood analytically. The corresponding equations for the low-risk group L read as

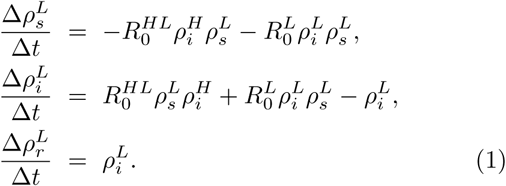

Here, 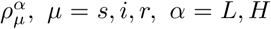, stands for the density of individuals within group *α* that are susceptible (*μ* = *s*), infectious (*μ* = *i*), and recovered (*μ* = *r*), respectively. The time-step of the discretized dynamics is denoted by Δ*t* and for SARS-CoV-2 roughly amounts to 5 days in real time [13]. The parameter 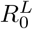 is the reproduction rate within group L, and the inter-group coupling 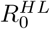 quantifies the transfer of infections from group H to group L. The corresponding equations for the high-risk group H are identical to Eqs. (1) upon exchanging *H* and *L* in all instances. The resulting parameters 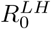 and 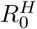 then denote the transfer of infections from L to H, and the effective reproduction rate within H, respectively. The goal of 2GROUPs is of course to keep 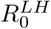 as low as possible, but it will still be non-zero in any realistic implementation.

It is important to notice that the reproduction numbers scale with the relative size of the groups: 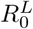 and 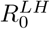 scale with the fraction of the population in the low-risk group while 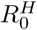 and 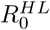 scale with the population fraction in the high-risk group. In a realistic scenario with the low-risk group being about five times as large as the high risk group this directly leads to a factor of five between 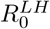 as compared to 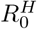 and *R*^*HL*^, as reflected in the parameters used for the simulations shown in Fig. 1. Moreover, in our example in Fig. 1, we use *R*^*LH*^ = 0.5 which corresponds to an additional three-fold reduction of contacts involving individuals from group H, compared to the already mitigated values (for SARS-CoV-2) within group L of 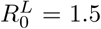 This, for the high-risk group, amounts to a rather strict isolation at a level of hard lockdowns in Europe during spring 2020, and even similar to the level reached during the lockdown in Wuhan [14].

To gain analytical insight, and motivated by the afore-mentioned scaling of parameters with relative group size, within a first approximation we neglect both 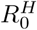 and 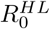, while keeping 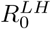 and 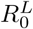 finite. This leads to a uni-directional decoupling of the two groups (note that H no longer appears in Eqs. (1) for the group L), which allows us to get an intuitive feeling for the synchronization of the two groups (see Fig. 2(a)). Corrections arising from restoring the neglected couplings are discussed further below. Roughly speaking, this only adds additional channels of infection compared to the simplifying approximation, and at least does not make the situation more favorable for the H group. Within our approximation, we may first solve Eqs. (1) for group L and then plug the time-dependent solution 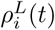 into the corresponding equations for *H*. Putting Δ*t* = 1, i.e. measuring time in units of Δ*t*, we thus derive

**FIG. 2:**
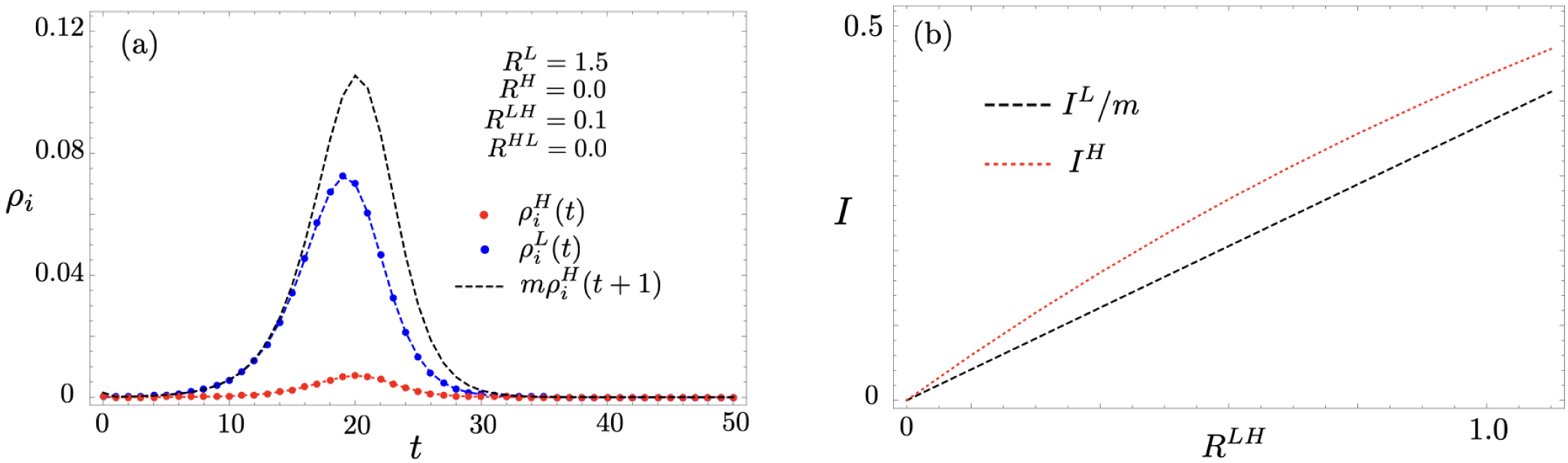
Instability at weak inter-group coupling (a) and total infection during the epidemic (b), illustrating Eq. (5) for various inter-group coupling values 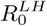 (evaluated at 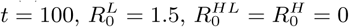 for the initial condition 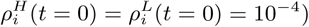).

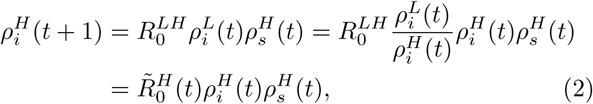

where we have defined 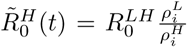 as an effective time-dependent reproduction rate for group *H*. Eq. (2) formally resembles a time-step in a simple epidemic dynamics of a single group but with 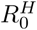 replaced by the time-dependent 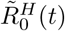. Importantly, since 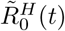 is pro-portional to the quotient 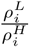, a much lower density of infected in the H group (the main goal of 2GROUPS) *increases* the epidemic growth within H – a very undesirable but unavoidable effect of the non-linear coupling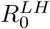. During the initial exponential growth of infections in L, a stable situation characterized by a time-independent 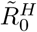 only occurs if the condition

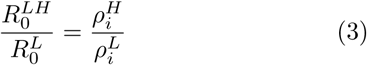

is satisfied. Then, the dynamics of 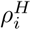 simply follows the dynamics of 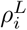 with a delay of a single time step and an overall reduction in amplitude, effectively dividing it by

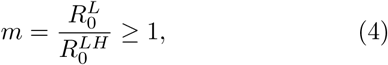

i.e. 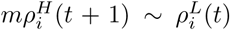 (cf. Eq. (3)). Close to the peak and during the downward slope of the dynamics, the delay causes 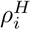 to grow even slightly above 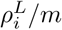 (see Fig. 1). Hence, a simple estimate for the overall number of infected *I*^*H*^ within the high-risk group is given by

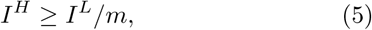

where *I*^*L*^ denotes the overall number of infected within the low-risk group during the epidemic (see Fig. 2(b)). These general results are illustrated in Figs. 1 and 2(a). In fact, a stronger bound holds throughout the relevant parts of the epidemic, namely that 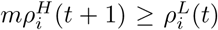 (cf. the black dashed and blue solid curves in Fig. 1). We stress that Fig. 1 shows data on the full model (1) with finite parameter values 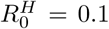 and 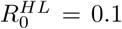, and the good agreement with our analytical predictions based on neglecting those couplings thus corroborates the robustness of our analytical picture over a wider parameter range.

More generally, we find that an undesirable outbreak within the high-risk group that, apart from a delay by one time-step, is qualitatively similar to the behavior of the low-risk group occurs as soon as 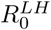 is non-zero, even for very small values (see Fig. 2(a)). Furthermore, increasing 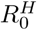 and 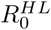 is never found to reduce 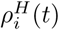 (or *I*^*H*^ for that matter). Hence, our analytical picture for 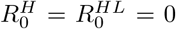 may be seen as an optimistic lower bound for the infections within the high-risk group.

## Concluding remarks

In this work, we have shown that the efficiency of a stratified epidemic strategy dividing the population into a low- and a high-risk group is drastically limited by synchronization effects occurring for any finite coupling between the groups. Specifically, the most optimistic hope to maintain a significantly slower effective reproduction rate for the high-risk group as compared to the low-risk group is largely ruled out. We have explicitly demonstrated this analytically in SIR-based 2GROUP models that give a coarse grained mean-field picture, noting that SIR models are microscopically more accurate for the spread of e.g. influenza viruses than for SARS-CoV-2, as clusters and superspreading events play an important role for the latter. Furthermore, certain quantitative aspects such as the delay in infections between the two groups may differ significantly due to finer structures of a real society not captured by our simple modelling [15]. Finally, we note that our model assumes perfect immunity upon recovery. That may be a reasonable approximation during a specific wave of the epidemic, and deviations from this assumption can for sure only make the situation worse for both groups. Despite our minimal modeling, we would find it very surprising, if the qualitative effects revealed in our present work, in particular the impossibility of sustaining a selective containment for a risk-group only, could not be clearly identified in more microscopic modelling scenarios. Along these lines, we hope that our findings stimulate future efforts to analyze more detailed COVID-19 specific models from a viewpoint of epidemic curves corresponding to different groups, including effects such as partial immunity, time-dependent strategies, imperfect vaccinations, and relaxing NPIs.

As a matter of fact, a sort of 2GROUPs strategy has been applied in Sweden for the first 7 months of the COVID-19 pandemic. While this has certainly contributed to keeping the fatalities lower than in an entirely unmitigated scenario, the comparably high fatality rates in Sweden may serve as a practical example of how hard it is to selectively protect high-risk groups [16, 17]. While insufficient security measures naturally play a major role here, we note that the outcome is in agreement with our general analysis: the substantial impact on the high-risk group has been reflecting the high spread in society at large. This stands in contrast to the neighboring countries that have applied a containment strategy directed to the society at large. Even without similarly stringent restrictions for the risk groups, this has resulted in a per capita fatality rate roughly an order of magnitude lower than in Sweden. This indicates that synchronization phenomena as the ones revealed in our present study might be more universal for epidemic dynamics, as long as infections occur between individuals (as opposed to disease spreading via agents such as mosquitos).

## Data Availability

All data and source code is available from the authors upon request.

## Acknowledgments

We would like to thank Marcus Carlsson, Thors Hans Hansson and Jan LÖtvall for useful comments and discussions. EJB acknowledges the Science Forum Covid-19 (https://vetcov19.se/en/) for numerous discussions on epidemiology and COVID-19. EJB is supported by the Swedish Research Council (VR) and the Wallenberg Academy Fellows program of the Knut and Alice Wallenberg Foundation.

